# Interaction Between Exercise Intensity and Sedentary Behavior on VO_2_max Change in Patients with De Novo Parkinson Disease: The Study in Parkinson Disease of Exercise (SPARX) Phase 2 Randomized Clinical Trial

**DOI:** 10.64898/2026.09.30.26364437

**Authors:** Byung Joon Pae, Tom S. Novak, Garett J. Griffith, Charity G. Patterson, Madeleine E. Hackney, Cory L. Christiansen, Daniel M. Corcos, Joe R. Nocera

## Abstract

**Background:** Parkinson disease (PD) is associated with increased sedentary time and reduced cardiorespiratory fitness, making exercise an important component of disease management. Although exercise improves cardiorespiratory fitness, individuals with PD vary substantially in their response to exercise. Sedentary time may contribute to this variability, yet whether sedentary time modifies the response to intervention and varying exercise intensities within an intervention is unknown. We examined whether sedentary time interacted with 6 months of moderate-intensity or high-intensity aerobic exercise on maximal oxygen consumption (VO₂max) in individuals with PD.

**Methods:** This secondary analysis used data from the SPARX phase 2 randomized clinical trial including 128 de novo PD patients assigned to high- or moderate-intensity treadmill exercise, or usual care for 6 months. Interactions between randomized group and post-randomization 6-months average sedentary time were assessed for associations with 6-month change in VO_2_max by using linear regression models corresponding to Analysis of Covariance (ANCOVA) models with interaction.

**Results:** The interaction between exercise intensity and sedentary behavior on 6-month change in VO_2_max in the moderate-intensity group compared with usual care was not significant (β: -1.78, 95% CI: -3.76 to 0.20, p-value: 0.08), although the sensitivity analysis restricted to complete cases showed a significant interaction where each 10% increase in 6-months average sedentary time was associated with a 2.51 mL/kg/min decrease in VO_2_max change in the moderate-intensity group compared with usual care (β: -2.51, 95% CI: -4.53 to -0.48, p-value: 0.02). There was no significant interaction between exercise intensity and sedentary behavior on 6-month change in VO_2_max in the high-intensity group compared with usual care (β: -1.46, 95% CI: -4.74 to 1.83, p-value: 0.38). The sensitivity analysis showed similar results.

**Conclusions:** These hypothesis-generating, preliminary findings suggest that sedentary time may modify the response to exercise among individuals participating in moderate-intensity aerobic exercise. Future adequately powered randomized controlled trials are needed to confirm these findings and determine whether sedentary behavior should be considered when prescribing exercise for individuals with PD.

## Introduction

Parkinson disease (PD) is the second most prevalent neurodegenerative disease, with global prevalence and incidence continuing to increase at an alarming rate.^1^ The Global Burden of Disease study estimated a global annual percentage change of 1.52% and 1.09% between 1990 and 2021 for age-standardized prevalence and incidence of PD, respectively.^1^ Considering the impact of PD on disability and quality of life,^2, 3, 4^ it is essential to identify and optimize interventions that can ameliorate health-related outcomes among PD patients.

Aerobic exercise is an established strategy for improving cardiorespiratory fitness in people with PD.^5, 6, 7^ However, limited evidence exists regarding whether the physiological benefits of exercise intensity depend on sedentary behavior during non-exercising time. Importantly, it has been suggested that individuals may compensate for exercise training by altering their behavior outside of exercise sessions, including increasing sedentary time.^8^ Conversely, emerging evidence suggests that reducing sedentary behavior in addition to engaging in exercise may provide benefits beyond those achieved with exercise alone.^9^ One potential mechanism is that exercise induces transient metabolic and molecular adaptations in skeletal muscle, including enhanced glucose uptake, insulin sensitivity, and oxidative metabolism,^10^ whereas high levels of sedentary behavior between exercise sessions may limit the maintenance of exercise-induced metabolic adaptations.^11^ Thus, sedentary behavior may represent an important behavioral context that influences physiological adaptation to exercise. Investigating whether sedentary time modifies the effects of exercise intensity during an exercise intervention could provide insight into variability in exercise response and inform more personalized exercise interventions for people with PD.

This study is a secondary analysis of the SPARX trial, which demonstrated that high-intensity aerobic exercise produced a significantly greater 6-month improvement in VO₂max compared with usual care, whereas moderate-intensity aerobic exercise did not produce a significantly greater 6-month improvement in VO₂max compared with usual care.^6^ This secondary analysis examined whether sedentary time modified the effects of high- and moderate-intensity aerobic exercise on 6-month changes in VO₂max. We hypothesize that, among de novo PD patients, greater amounts of time in sedentary behavior during non-exercise time will be associated with attenuated improvements in VO_2_max in both the high- and moderate-intensity exercise groups relative to usual care.

## Methods

### Study Participants

Data from the Study in Parkinson Disease of Exercise (SPARX) phase 2 randomized clinical trial were used for this secondary analysis. SPARX was a randomized clinical trial of de novo PD patients that investigated the effects of high- and moderate-intensity treadmill exercise compared with usual care (a wait-list control group).^6^ Individuals were enrolled from outpatient and community-based clinics between May 1, 2012, through November 30, 2015 in Chicago, Illinois; Denver, Colorado; and Pittsburgh, Pennsylvania.^6^ Participants were between 40 and 80 years of age, had Hoehn and Yahr stage 1-2 disease, had been diagnosed within the past 5 years, were not engaging in moderate-intensity exercise more than three times per week, and were not anticipated to require dopaminergic medication within the following 6 months. A total of 128 participants were randomized into high-intensity treadmill exercise, moderate-intensity treadmill exercise, or usual care (Figure 1). All participants provided written informed consent, and all data were deidentified. The protocol was approved by institutional review boards of all participating sites, and the trial was registered at clinicaltrials.gov (Identifier: NCT01506479).

**Figure 1.**
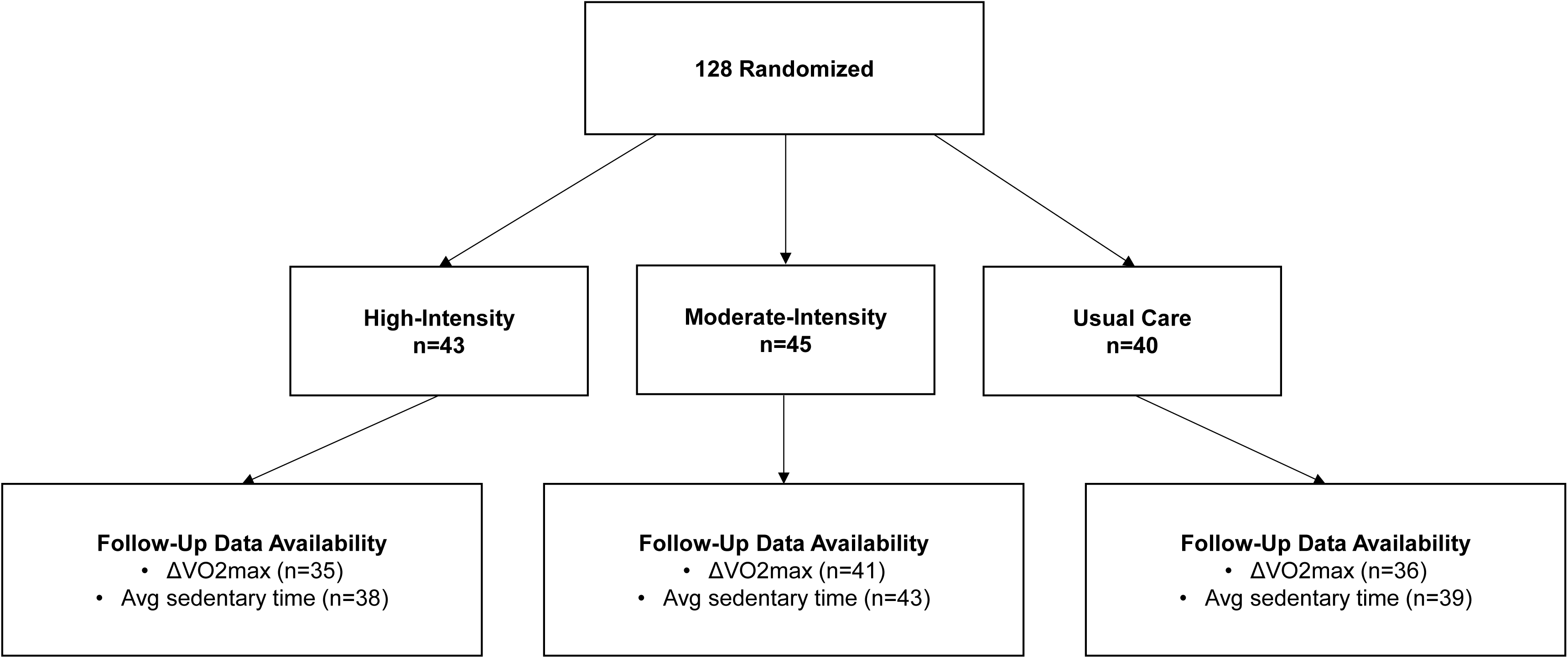
Flowchart of the study participants. VO_2_max, maximal oxygen consumption; AVG, average; HR, heart rate.

## Exercise Interventions

Treadmill exercise was performed for 26 weeks (6 months) and consisted of 5 to 10 minutes of warm-up, 30 minutes of treadmill exercise at the target heart rate, and 5 to 10 minutes of cool down. Regarding target heart rate, cardiopulmonary exercise tests (CPET) were performed to measure directly maximal heart rates. The high-intensity group performed treadmill exercise at 80%-85% of maximum heart rate, and the moderate-intensity group performed treadmill exercise at 60%-65% of maximum heart rate. The usual care group was asked to maintain exercise habits. Participants were asked to perform treadmill exercise 4 days per week. The first 8 weeks served as a ramp-up period during which exercise frequency and intensity were slowly increased to reach target heart rate levels. During all exercise sessions, heart rate monitors were used to record the intensity of treadmill exercise. For participants who initiated chronotropic medications, exercise intensity was monitored by using rating of perceived exertion (RPE). All exercise sessions occurring within the first 2 weeks were performed under on-site supervision, and participants subsequently visited the respective study site twice-monthly for supervised sessions and provided heart rate data to download. Monthly conference calls with study coordinators were implemented to maintain protocol fidelity.

## Sedentary Behavior

Sedentary time was extracted using tri-axial, waist-worn accelerometers (ActiGraph GT3X [+ and BT]; ActiGraph, Pensacola, FL).^12^ Acceleration data were sampled at 60 Hz and processed using commercially available software (ActiLife 5/6; ActiGraph).^12^ The “Sedentary” classification from the ActiLife software output was used to define sedentary minutes, which were any 60-second epoch with less than 100 acceleration vector counts per minute. Participants wore the accelerometer for up to 10-day periods, conducted once per month over 6 months. Average daily sedentary time was calculated as mean daily proportion of ActiGraph wear time spent in sedentary behavior. For each month, average daily sedentary proportions were calculated over the wear period to obtain a month-specific estimate. These month-specific estimates were then averaged across the six monitoring periods to derive a participant-level mean sedentary time over 6 months. The corresponding 6-months average ActiGraph wear time (minutes/day) was derived using the same procedure. To prevent selection bias and preserve the 6-months average construct, no minimum wear time or valid wear day criteria were applied. As PD patients with more severe symptoms may have greater difficulty complying with wearable devices,^13^ imposing such criteria may retain healthier observations. Furthermore, such criteria may drop any of the 6 monitoring periods and break the 6-months average construct, preventing calculation of the intended 6-months average sedentary behavior across all 6 monitoring periods.

## Outcome

The primary outcome of this study was 6-month change in VO_2_max (follow-up - baseline). During supervised maximal graded exercise tests, expired gas analyses were sampled to measure oxygen consumption and carbon dioxide production via indirect calorimetry (ParvoMedics TrueOne 2400, Salt Lake City, UT for Chicago and Denver; AEI Technologies, MOXUS, Bastrop, TX for Pittsburgh). The test consisted of a warm-up period on the treadmill to identify the walking speed and grade that generates a heart rate corresponding to 65%-70% of the age-predicted maximum heart rate, calculated as 220 - age,^14^ followed by a brief rest period to initiate the indirect calorimetry. The test was subsequently initiated at the appropriate walking speed and 0% grade. Thereafter, treadmill grade was increased by 2% every 2 minutes until volitional exhaustion or test termination due to abnormal responses.

## Statistical Analysis

Baseline characteristics of study participants were summarized by randomized groups (high-intensity, moderate-intensity, and usual care). Continuous variables were reported as mean (SD), and categorical variables were reported as count (%). As a primary objective of this study, planned interaction effects were assessed using linear regression models including randomized group, sedentary behavior, and their interaction term, corresponding to Analysis of Covariance (ANCOVA) models with interaction. Given the hypothesis-generating nature of this secondary study, a parsimonious modelling approach was utilized to optimize statistical power. Because the 6-months average sedentary time, including 6-months average accelerometer wear time, represented post-randomization behavior, additional linear regression models equivalent to One-Way Analysis of Variance (ANOVA) were conducted to test the overall F-statistic for any significant differences among the randomized groups. Two-sided p-value < 0.05 was considered statistically significant.

For all analyses, randomized group was treated as a categorical variable with usual care serving as the reference group. Average daily sedentary time was treated as a continuous variable. For assessing interaction effects, daily sedentary time was mean-centered and rescaled to represent per 10% increase. To visualize the interaction between exercise intensity and sedentary behavior on 6-month change in VO_2_max, predicted means were plotted by tertiles of 6-months average sedentary time.

Primary analyses incorporated multiple imputation for the follow-up VO_2_max and 6-months average sedentary time with an intention-to-treat approach. Specifically, Multiple Imputation by Chained Equations (MICE) was used.^15^ Because loss to follow-up differed by randomized group, the complete-case data were considered potentially biased. Continuous variables were thus imputed using linear regression models without predictive mean matching to avoid restricting imputed values to the observed range of the sample.^16^ Results from MICE were pooled using Rubin’s rule to derive pooled parameter estimates.^15, 17^

Sensitivity analyses addressing missing data were performed using complete cases. Additional sensitivity analyses evaluated robustness to minimum wear time threshold and varying number of monthly monitoring periods across both multiple imputation and complete-case data. These analyses utilized the same ANCOVA-equivalent models with interaction but were applied a minimum wear time threshold of 10 hours/day to daily accelerometer data and incorporated weighted least squares, weighting by each participant’s number of monthly monitoring periods. All statistical analyses were performed using SAS software (Version 9.4; SAS Institute, Cary, NC, US).

## Results

### Baseline Characteristics

A total of 128 participants were randomized into high-intensity treadmill exercise (n=43, 53.5% age ≥ 65 years, 51.2% males, 90.7% whites), moderate-intensity treadmill exercise (n=45, 44.4% age ≥ 65 years, 60.0% males, 88.9% whites), or usual care (n=40, 55.0% age ≥ 65 years, 60.0% males, 90.0% whites). Baseline characteristics of the three randomized groups were well balanced (Table 1).

**Table 1.** Baseline Characteristics of Study Participants

|  | **High-Intensity**<br>**Group (n=43)** | **Moderate-**<br>**Intensity Group**<br>**(n=45)** | **Usual Care**<br>**Group (n=40)** |
| --- | --- | --- | --- |
| Age ≥ 65 years, n (%) | 23 (53.5) | 20 (44.4) | 22 (55.0) |
| Male, n (%) | 22 (51.2) | 27 (60.0) | 24 (60.0) |
| Race, n (%) |  |  |  |
| White | 39 (90.7) | 40 (88.9) | 36 (90.0) |
| Non-White | 4 (9.3) | 5 (11.1) | 4 (10.0) |
| BMI ≥ 25, n (%) | 25 (58.1) | 28 (62.2) | 26 (65.0) |
| MDS-UPDRS |  |  |  |
| Part 1 score | 5.8 (4.2) | 5.2 (3.5) | 4.9 (3.8) |
| Part 2 score | 5.5 (4.0) | 5.5 (4.1) | 5.5 (3.8) |
| Part 3 score | 22.4 (9.0) | 21.1 (8.9) | 21.1 (9.1) |
| Part 4 score | 0.1 (0.3) | 0.2 (0.7) | 0.2 (0.5) |
| VO<sub>2</sub>max, mL/kg/min | 22.9 (6.1) | 24.4 (7.0) | 24.0 (4.4) |

Data are presented as n (%) for categorical variables and mean (SD) for continuous variables unless otherwise stated.
BMI, body mass index; MDS-UPDRS, movement disorder society-unified Parkinson's disease rating scale; VO<sub>2</sub>max, maximal oxygen consumption.

## Sedentary Behavior

There were no differences in 6-months average sedentary time across the three randomized groups (Table 2). Sensitivity analyses restricted to complete cases yielded similar results (Table S1).

**Table 2.** Comparison of 6-Months Average Sedentary Time by Randomized Group in the Multiple Imputation Data (n=128)

|  | **High-Intensity Mean (SE)** | **Moderate-Intensity Mean (SE)** | **Usual Care Mean (SE)** | **F-Statistic** | **P-value** |
| --- | --- | --- | --- | --- | --- |
| 6-Months Average Sedentary Time, % | 53.8 (2.5) | 56.0 (1.4) | 56.0 (1.5) | 0.46 | 0.63 |

One-way ANOVA was used to compare 6-months average sedentary time across randomized groups.

Regarding the 6-months average accelerometer wear time (minutes/day), there were no significant differences across the three randomized groups among complete cases. All groups had a 6-months average accelerometer wear time of at least 766 minutes/day (12.7 hours/day) (Table S2).

## Changes in VO_2_max

During the 6-month exercise intervention period, the average change in VO_2_max was 2.6 ± 5.4, - 0.1 ± 4.4, and -1.2 ± 2.6 mL/kg/min for the high-intensity, moderate-intensity, and usual care group, respectively (Table 3).

**Table 3.** 6-Month Change in VO_2_max by Randomized Group in the Multiple Imputation Data (n=128)

|  | **High-Intensity**<br>**Mean (SD)** | **Moderate-Intensity**<br>**Mean (SD)** | **Usual Care**<br>**Mean (SD)** |
| --- | --- | --- | --- |
| 6-Month Change in<br>VO<sub>2</sub>max, mL/kg/min | 2.6 (5.4) | -0.1 (4.4) | -1.2 (2.6) |

VO<sub>2</sub>max, maximal oxygen consumption.

## Interaction Effects between Exercise Intensity and Sedentary Behavior

There was no interaction between exercise intensity and sedentary behavior on 6-month change in VO_2_max for the high-intensity group (β: -1.46, 95% CI: -4.74 to 1.83, p-value: 0.38) or the moderate-intensity group (β: -1.78, 95% CI: -3.76 to 0.20, p-value: 0.08) compared with usual care (Table 4). From the same model, the high-intensity group showed greater 6-month change in VO_2_max compared with usual care at the mean value of sedentary behavior (β: 3.52, 95% CI: 1.63 to 5.42, p-value: < 0.001) (Table 4). Figure 2 presents the interaction between exercise intensity and 6-months average sedentary time on 6-month change in VO_2_max. In sensitivity analyses restricted to complete cases, each 10% increase in 6-months average sedentary time was associated with a 2.51 mL/kg/min decrease in VO_2_max change in the moderate-intensity group compared with the usual care group (β: -2.51, 95% CI: -4.53 to -0.48, p-value: 0.02) (Table S3). There was no interaction effect for the high-intensity group among complete cases (β: -0.31, 95% CI: -2.32 to 1.71, p-value: 0.76) (Table S3). Similar results were observed in sensitivity analyses applying the minimum wear time threshold (≥10 hours/day) and weighted least squares across both multiple imputation and complete-cases data (Tables S4-S5). Overall patterns were consistent across analyses.

**Figure 2.**
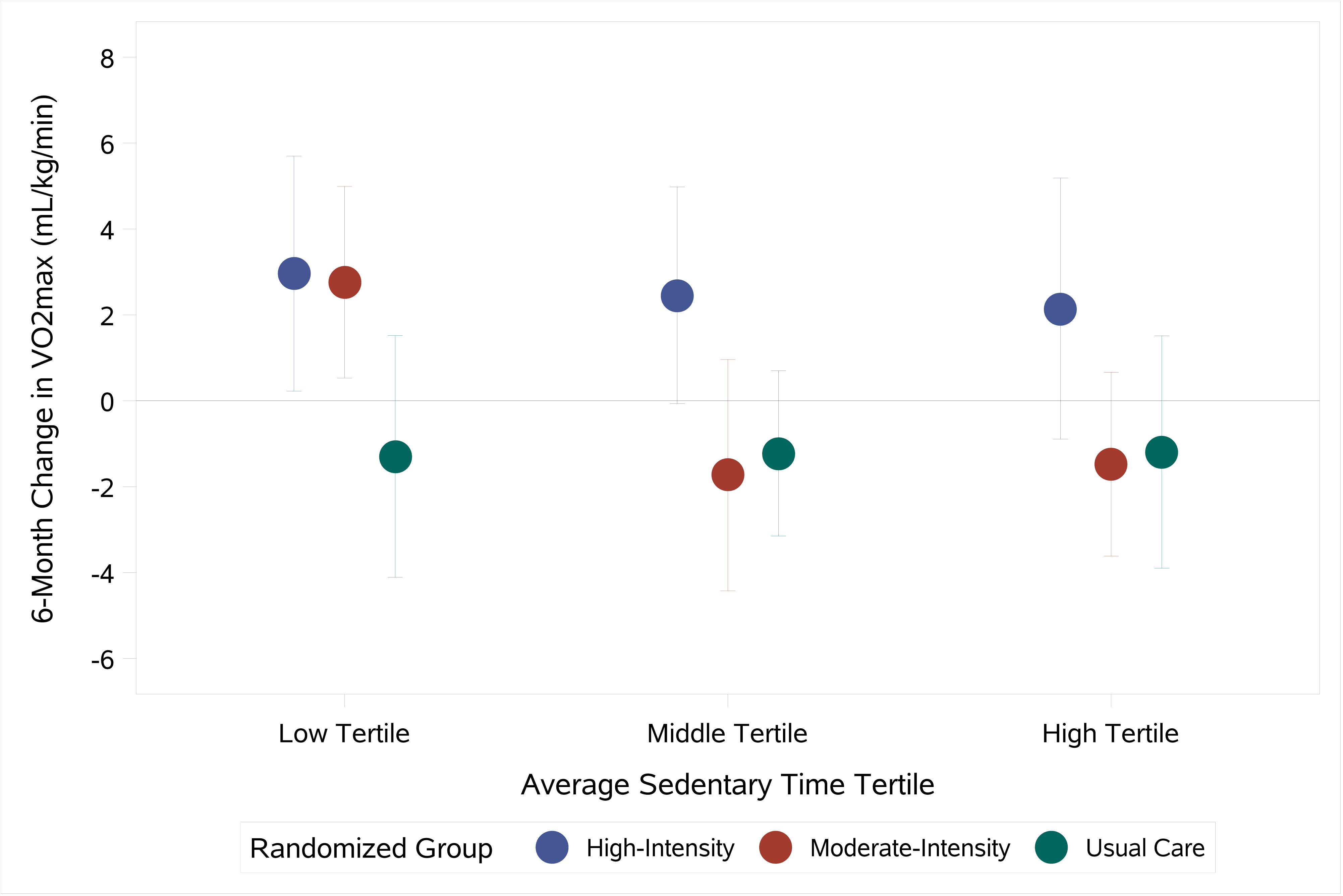
Interaction between exercise intensity and tertiles of 6-months average sedentary time on 6-month change in VO_2_max in the multiple imputation data (n=128). Data shown are predicted means derived from the ANCOVA model including randomized group, tertiles of 6- months average sedentary time, and the interaction term, presented for visualization purposes. Error bars represent 95% confidence intervals. Sample sizes: High-Intensity/Low Tertile (n=17), High-Intensity/Middle Tertile (n=12), High-Intensity/High Tertile (n=14), Moderate- Intensity/Low Tertile (n=15), Moderate-Intensity/Middle Tertile (n=12), Moderate- Intensity/High Tertile (n=18), Usual Care/Low Tertile (n=10), Usual Care/Middle Tertile (n=19), Usual Care/High Tertile (n=11). VO_2_max, maximal oxygen consumption; ANCOVA, analysis of covariance.

**Table 4.** Interaction between Exercise Intensity and 6-Months Average Sedentary Time on 6- Month Change in VO_2_max in the Multiple Imputation Data (n=128)

|  | Main Effect |  | Interaction |  |
| --- | --- | --- | --- | --- |
| | $\beta$ (95% CI) | *P*-value | $\beta$ (95% CI) | *P*-value |
| High-Intensity | 3.52 (1.63, 5.42) | <0.001 | -1.46 (-4.74, 1.83) | 0.38 |
| Moderate-Intensity | 1.24 (-0.53, 3.01) | 0.17 | -1.78 (-3.76, 0.20) | 0.08 |
| Usual Care | REF | REF | REF | REF |

6-months average sedentary time was mean-centered and rescaled to represent per 10% increase. Main effect represents differences in high- and moderate-intensity groups compared with usual care at the mean value of 6-months average sedentary time. Model included randomized group, 6-months average sedentary time, and the interaction term. VO<sub>2</sub>max, maximal oxygen consumption; CI, confidence interval.

## Discussion

The current, secondary analysis of the SPARX trial tested for interaction effects between exercise intensity and sedentary time on 6-month change in VO_2_max in de novo PD patients. In this analysis, greater sedentary behavior was associated with a less favorable change in cardiorespiratory fitness among participants randomized to moderate-intensity exercise compared with usual care, although this association was evident only in the complete-case sensitivity analysis. Importantly, the observed association was not simply between sedentary behavior and baseline or absolute cardiorespiratory fitness, rather, greater sedentary time was associated with a smaller improvement in VO₂max following moderate-intensity exercise relative to usual care. The magnitude of this association was notable, with each 10% increase in sedentary time corresponding to a 2.51 mL/kg/min smaller improvement in VO₂max. Although the clinical significance of this difference cannot be established from this exploratory analysis, it suggests that differences in daily sedentary behavior could meaningfully influence the physiological benefits obtained from moderate-intensity exercise and carry important prognostic implications.^18^ This finding raises the possibility that sedentary behavior may influence the capacity to adapt to a moderate exercise stimulus rather than simply reflecting differences in baseline fitness. In contrast, no significant interaction between sedentary behavior and exercise intensity was observed for the high-intensity exercise group. Importantly, the interaction did not reach statistical significance in the primary analysis (p-value: 0.08), and the association became significant only when analyses were restricted to complete cases. Thus, the finding should be interpreted as preliminary rather than confirmatory. The consistency in the direction of the effect across analyses, along with substantial overlap in 95% confidence intervals, provides a rationale for further investigation.

The finding suggesting no relationship between high-intensity exercise and sedentary behavior is consistent with previous studies.^19, 20^ One potential biological explanation for the apparent difference between exercise intensities involves the regulation of mitochondrial biogenesis and oxidative metabolism. Sedentary behavior has been associated with reduced PGC-1α signaling, whereas higher-intensity exercise produces a stronger stimulus for PGC-1α and mitochondrial adaptations.^21, 22, 23^ It is therefore possible that the larger physiological stimulus provided by high-intensity exercise may be less susceptible to the metabolic effects associated with sedentary behavior. This mechanism is speculative, however, as PGC-1α and other molecular markers were not measured in SPARX.

While these exploratory analyses are not definitive, there are several key clinical implications related to structuring prescriptive exercise programs for VO_2_max improvements in PD patients. Although high-intensity exercise may provide robust physiological benefits despite greater sedentary behavior, our findings raise the possibility that the benefits of moderate-intensity exercise may be more sensitive to sedentary behavior. Thus, reducing sedentary time may represent an important complement to moderate-intensity exercise, particularly for individuals with PD who are unable to tolerate or participate in high-intensity exercise. More broadly, these findings raise the possibility that exercise responsiveness in PD is influenced not only by the characteristics of the prescribed exercise stimulus but also by behaviors occurring outside of exercise sessions, particularly among PD patients participating in moderate-intensity exercise.

This study has several limitations that must be addressed. Because this secondary, exploratory study used data from SPARX, which was a phase 2 trial not adequately powered to detect interaction effects, the findings should be considered exploratory and hypothesis-generating.^24^ Additionally, sedentary behavior was calculated as the average sedentary time during the 6-month exercise intervention period and was not randomized; therefore, interaction estimates should not be interpreted causally. However, 6-months average sedentary time did not differ significantly across randomized groups, supporting the evaluation that sedentary time during the intervention may differentially relate to VO_2_max response by exercise intensity assignment. Future studies should attempt to investigate the interactions between exercise intensity and sedentary behavior on health-related outcomes among PD patients using well powered trials that incorporate randomized strategies.

## Conclusion

The findings of this secondary, exploratory study have demonstrated that, among de novo PD patients, greater sedentary behavior was not associated with attenuated improvements in VO_2_max in the high-intensity exercise group relative to usual care but showed a non-significant trend toward blunting the effect of moderate-intensity exercise, which was only significant in the complete-case data. These findings warrant future studies to confirm the current findings and investigate whether different sedentary behavior contexts may be considered for the prescription of structured exercise programs for PD patients. Furthermore, whether interventions utilizing moderate-intensity protocols may consider incorporating targeted behavioral strategies to reduce prolonged sedentary behaviors to maximize cardiorespiratory benefits should be investigated. Given the underpowered nature of this secondary, exploratory study, findings must be interpreted as hypothesis-generating. Further research using adequately powered randomized controlled designs is warranted to better understand how exercise intensity interacts with behavioral factors such as sedentary behavior on health-related outcomes in patients with PD.

## Data Availability

The original SPARX data was shared with NINDS in May, 2017, and all requests should be sent to NINDS

## Acknowledgements

We thank the participants and investigators of the SPARX study for their significant and important contributions.

## Supplemental Material

Tables S1-S5

