## Supplemental Material for "Interaction Between Exercise Intensity and Sedentary Behavior on VO_2_max Change in Patients with De Novo Parkinson Disease: The Study in Parkinson Disease of Exercise (SPARX) Phase 2 Randomized Clinical Trial"

**Table S1**. Comparison of 6-Months Average Sedentary Time by Randomized Group among Complete Cases (n=120)

|  | **High-Intensity**  **Mean (SD)** | **Moderate-Intensity**  **Mean (SD)** | **Usual Care**  **Mean (SD)** | **F-Statistic** | ***P*-value** |
| --- | --- | --- | --- | --- | --- |
| 6-Months Average Sedentary Time, % | 55.5 (10.3) | 55.6 (8.8) | 56.2 (9.6) | 0.05 | 0.95 |

One-way ANOVA was used to compare 6-months average sedentary time across randomized groups.

**Table S2**. Comparison of 6-Months Average Accelerometer Wear Time by Randomized Group among Complete Cases (n=120)

|  | **High-Intensity**  **Mean (SD)** | **Moderate-Intensity**  **Mean (SD)** | **Usual Care**  **Mean (SD)** | **F-Statistic** | ***P*-value** |
| --- | --- | --- | --- | --- | --- |
| 6-Months Average Accelerometer Wear Time, min/day | 766.1 (109.5) | 787.4 (97.0) | 769.4 (104.4) | 0.50 | 0.61 |

One-way ANOVA was used to compare 6-months average accelerometer wear time across randomized groups.

**Table S3**. Interaction between Exercise Intensity and 6-Months Average Sedentary Time on 6-Month Change in VO_2_max among Complete Cases (n=108)

|  | **Main Effect** | | **Interaction** | |
| --- | --- | --- | --- | --- |
|  | **β (95% CI)** | ***P*-value** | **β (95% CI)** | ***P*-value** |
| High-Intensity | 3.32 (1.70, 4.95) | **<0.001** | -0.31 (-2.32, 1.71) | 0.76 |
| Moderate-Intensity | 1.54 (0.00, 3.07) | 0.05 | -2.51 (-4.53, -0.48) | **0.02** |
| Usual Care | REF | REF | REF | REF |

6-months average sedentary time was mean-centered and rescaled to represent per 10% increase.

Main effect represents differences in high- and moderate-intensity groups compared with usual care at the mean value of 6-months average sedentary time.

Model included randomized group, 6-months average sedentary time, and the interaction term.

VO_2_max, maximal oxygen consumption; CI, confidence interval.

**Table S4**. Interaction between Exercise Intensity and 6-Months Average Sedentary Time on 6-Month Change in VO_2_max in the Multiple Imputation Data, Applying Minimum Wear Time Threshold (≥10 hours/day) and Weighted by Number of Monthly Monitoring Periods (n=128)

|  | **Main Effect** | | **Interaction** | |
| --- | --- | --- | --- | --- |
|  | **β (95% CI)** | ***P*-value** | **β (95% CI)** | ***P*-value** |
| High-Intensity | 3.71 (1.58, 5.84) | **<0.001** | -1.79 (-5.88, 2.31) | 0.39 |
| Moderate-Intensity | 1.50 (-0.37, 3.36) | 0.12 | -1.91 (-4.06, 0.23) | 0.08 |
| Usual Care | REF | REF | REF | REF |

6-months average sedentary time was mean-centered and rescaled to represent per 10% increase.

Main effect represents differences in high- and moderate-intensity groups compared with usual care at the mean value of 6-months average sedentary time.

Model included randomized group, 6-months average sedentary time, and the interaction term.

VO2max, maximal oxygen consumption; CI, confidence interval.

**Table S5**. Interaction between Exercise Intensity and 6-Months Average Sedentary Time on 6-Month Change in VO_2_max among Complete Cases, Applying Minimum Wear Time Threshold (≥10 hours/day) and Weighted by Number of Monthly Monitoring Periods (n=108)

|  | **Main Effect** | | **Interaction** | |
| --- | --- | --- | --- | --- |
|  | **β (95% CI)** | ***P*-value** | **β (95% CI)** | ***P*-value** |
| High-Intensity | 3.32 (1.69, 4.96) | **<0.001** | -0.39 (-2.36, 1.58) | 0.70 |
| Moderate-Intensity | 1.66 (0.12, 3.21) | **0.04** | -2.57 (-4.54, -0.59) | **0.01** |
| Usual Care | REF | REF | REF | REF |

6-months average sedentary time was mean-centered and rescaled to represent per 10% increase.

Main effect represents differences in high- and moderate-intensity groups compared with usual care at the mean value of 6-months average sedentary time.

Model included randomized group, 6-months average sedentary time, and the interaction term.

VO2max, maximal oxygen consumption; CI, confidence interval.
